# Clinical Performance of BD Veritor™ Assay Across SARS-CoV-2 Variants

**DOI:** 10.1101/2023.04.11.23288130

**Authors:** Karen Eckert, Sebastian Gutierrez, Brittany Knight, Lauren Cooper

**Author notes:** **To whom correspondence should be addressed** Lauren Cooper, PhD, MPH, D(ABMM), Senior Manager, Scientific Affairs, Becton, Dickinson and Company, BD Life Sciences – Integrated Diagnostic Solutions (IDS), 7 Loveton Circle, Sparks, MD 21152, USA.

## Abstract

**Background:** Different rates of morbidity, mortality, transmission, and immune escape are associated with various strains of the SARS-CoV-2 virus. With the emergence of new strains during seasonal outbreaks, ensuring that antigen-based immunoassays can detect SARS-CoV-2 infections across identified circulating viral variants is a crucial component of infection control efforts.

**Objective:** To validate the performance of the BD Veritor System for Rapid Detection of SARS-CoV-2 Assay (BD Veritor assay) to detect SARS-CoV-2 across variants of concern (VOC) and variants of interest (VOI).

**Methods:** Using the Illumina NextSeq 2000 Sequencer, viral sequencing was performed on prospectively collected, then frozen, SARS-CoV-2 RT-PCR positive nasal swabs stored in universal transport media (UTM). Specimens from symptomatic and asymptomatic individuals were included in the study. Using the information obtained from the sequencing analysis, the performance of the BD Veritor System assay was evaluated against the highly sensitive molecular RT-PCR Quidel Lyra SARS-CoV-2 assay for each variant.

**Results:** The resulting PPA was 97.4% (95% CI: 86.8, 99.5) for detection of SARS-CoV-2 across all variants identified by Next Generation Sequencing (i.e., WHO-labeled variants Alpha, Delta, Gamma, Iota, Lambda, as well as two other non-labeled variants), with a 100% PPA for five of the six variant labels identified.

**Conclusion:** The results demonstrate the robust performance of the BD Veritor assay in detecting SARS-CoV-2 in clinical nasal specimens in selected variants. As new variants emerge, additional studies will be beneficial to ensure the sustained performance of SARS-CoV-2 assays.

## INTRODUCTION

Since the outset of the SARS-CoV-2 pandemic, five VOC and two VOI have been reported in North America, that is, Alpha, Beta, Delta, Gamma, and Omicron as VOC, and Lambda and Mu as VOI.^1^ The predicted seasonal pattern of SARS-CoV-2 activity in the endemic environment^2,3^ coupled with the known propensity of RNA viruses to mutate^4^ will likely result in varying rates of morbidity, mortality, transmission, and immune escape for each variant. Accordingly, concerns are raised over the phenotypic characteristics of each variant and their subsequent detectability with antigen-based immunoassays.

Recently, efforts have been ongoing to trace the various lineages of the SARS-CoV-2 virus through its viral genome using sequencing techniques.^5,6^ Currently, the gold standard to characterize and identify variants is the use of Next Generation Sequencing (NGS), however, the performance of multiplex RT-PCR assays has also recently been evaluated for that purpose.^7-10^ Irrespective of the method selected, the accurate initial detection of positive SARS-CoV-2 samples remains the basis upon which variant detection can be performed. As such, it is essential that the performance of current assays be established in terms of their demonstrated ability to detect SARS-CoV-2, regardless of the causative variant. To this aim, we evaluated the performance of the BD Veritor System for Rapid Detection of SARS-CoV-2 (Becton, Dickinson, and Company; BD Life Sciences – Integrated Diagnostic Solutions, Sparks, MD, USA) across identified WHO variants.

## METHODS

### Demographics

Both symptomatic and asymptomatic individuals ≥18 years old were eligible to participate in the SARS-CoV-2 parent study from which the positive specimens for this investigation were acquired. Inclusion criteria for asymptomatic adults were a recent known exposure to SARS-CoV-2 or a positive SARS-CoV-2 test. For symptomatic individuals, an onset of symptoms consistent with SARS-CoV-2 within 5 days of enrollment or a positive SARS-CoV-2 test within 72 hours prior to enrollment were the inclusion criteria. Individuals unable to provide the minimum number of swabs required, with frequent or difficult to control nosebleeds within 14 days of study participation, and those whose health was likely to be jeopardize from involvement in the investigation were not eligible for enrollment.

### Specimen collection and processing

Deidentified SARS-CoV-2 RT-PCR-positive remnant specimens from a parent study were used in this investigation. The nasal specimens (swabs) were collected prospectively by healthcare providers across three centers in the US from May 11 to July 25, 2021, stored in 3mL UVT, frozen at -70°C, and shipped on dry ice to TriCore Reference Laboratories for reference testing and storage. Specimens were shipped on dry ice to MRIGlobal at a later date for viral sequencing. To perform the sequencing, RNA was extracted using the Qiagen QIAamp Viral RNA Mini Kit (Qiagen Life Sciences, Hilden, Germany) with extracted nucleic acid subsequently placed into an Illumina RNA Preparation with Enrichment kit (Illumina Inc., San Diego, California, US) for reverse transcription, tagmentation, and barcoding using IDT® for Illumina® Unique Dual Indexes. Target enrichment using the Illumina Respiratory Virus Oligo Panel (RVOP) V2 was then performed, and a dual-index paired-end 2 × 151 sequencing was conducted on the Illumina NextSeq 2000 Sequencer. The Illumina DRAGEN COVID Lineage app (V3.5.9) was used to perform the variant analysis. The performance of the BD Veritor System assay was evaluated against the highly sensitive molecular RT-PCR Lyra assay (Lyra SARS-CoV-2 Assay; Quidel Corporation, Athens, OH, USA) for each variant.

Approval from Advarra Institutional Review Board (Columbia, MD) was obtained prior to study initiation. The study was conducted according to the principles set forth by the Declaration of Helsinki and Good Clinical Practices, and this manuscript was developed following the standards for reporting accuracy studies (STARD) guidelines.^11^

### Data analysis

PPA calculated with 95% confidence intervals (CI) using the Wilson score method was the primary outcome measure. Discordant results on the RT-PCR were re-tested on the same assay and results from the second testing were used for analysis.

## RESULTS

A total of 39 specimens from individuals (21 females [53.8%] and 18 males [46.2%]) with a positive SARS-CoV-2 RT-PCR test result were evaluated. Although both symptomatic and asymptomatic individuals were eligible for participation in the parent study, all 39 specimens used for the sequencing component of the investigation were from symptomatic individuals as no specimens from asymptomatic participants tested positive by RT-PCR. The age distribution of the participants was as follows: 10.3% (4/39) were 18-24 years old, 66.6% (26/39) were 25-64 years old, and 23.1% (9/39) were ≥65 years of age. Hispanic/Latino participants accounted for 76.9% (30/39) of the enrollment. Of the 39 positive specimens, two had insufficient depth of coverage to make a high confidence lineage identification and were consequently excluded from subsequent analysis, resulting in 37 evaluable specimens for lineage stratification. No invalid results were recorded on the BD Veritor assay or RT-PCR Lyra assay for included specimens. An average of 3.12 days (SD 1.17) between enrollment time and symptom onset was reported, and four of the 39 participants (10.3%) were positive for SARS-CoV-2 at time of enrollment.

PPA across the individual variants was 100% for five of the six lineage categories, and 87.5% for the Gamma variant (**Table 1**). The average Ct score for all variants detected by Lyra was 21.37 (range: 19.54-23.16, Median: 21.10). Individual and mean Ct scores by variant are presented in **Figure** 1; individual Ct values ranged from 15.31 to 29.99, inclusively, across categories.

**Figure 1.**
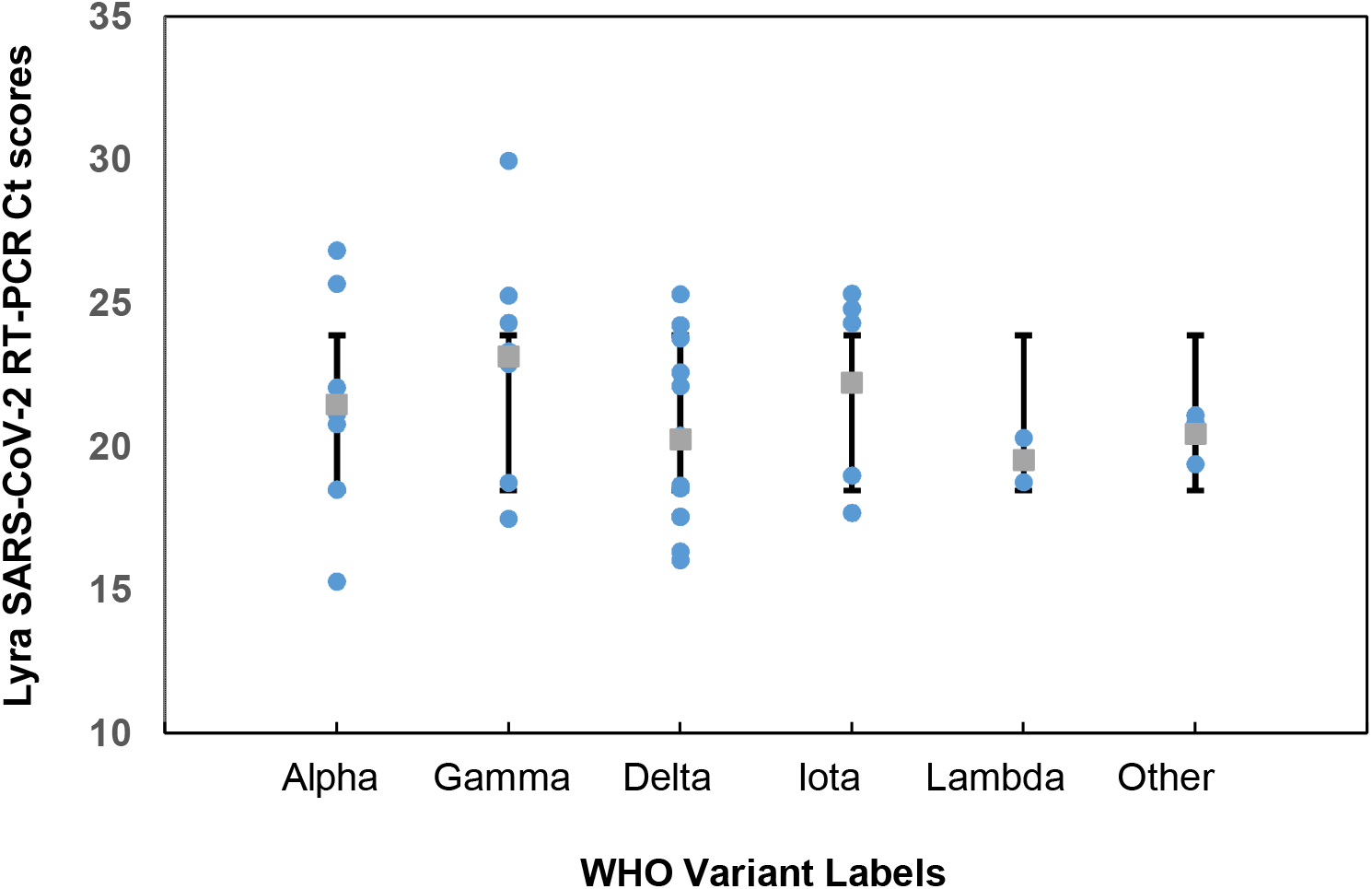
Individual plot of Lyra Ct scores by WHO-labeled variant. Individual positive results by variant are plotted according to the Ct score obtained with the Lyra SARS-CoV-2 RT-PCR assay for each specimen with blue circles indicate individual Lyra results and gray boxes showing the mean for each group with 95% CI.

**Table 1.** Assay Performance by Variant Stratification.

| **Variant** | **n/N Concordance** | **PPA (95% CI)** | **Lyra Average Ct Score** |
| --- | --- | --- | --- |
| Alpha | 7/7 | 100% (64.6, 100.0) | 21.48 |
| Gamma | 7/8 | 87.5% (52.9, 97.7) | 23.16 |
| Delta | 12/12 | 100% (75.5, 100.0) | 20.27 |
| Iota | 5/5 | 100% (56.5, 100.0) | 22.24 |
| Lambda | 2/2 | 100% (34.2, 100.0) | 19.54 |
| Other | 3/3 | 100% (43.8, 100.0) | 20.45 |
| **Total** | **36/37** | **97.4% (86.8, 99.5)** | **21.37** |

Ct: cycle threshold.

## DISCUSSION

The performance of an authorized rapid antigen test must be robust enough to allow the detection of positive samples, regardless of the SARS-CoV-2 virus lineage. Next generation genome sequencing for this study showed a 97.4% PPA (95% CI: 86.8, 99.5) across the 37 sequenced samples while performance across the individual variants was 100% for the *Alpha, Delta, Iota, Lambda* and *Other* categories (which included one B.1.637 and two XB lineage not labeled by WHO), and 87.5% (95% CI: 52.9, 97.7) for *Gamma*. Whereas a relatively limited sample size was used, the confirmed detectability of different variants by the BD Veritor System assay in our study aligns with results from other internal BD validation studies that used live or heat inactivated isolates with the BD Veritor Plus Analyzer to confirm detectability of SARS-CoV-2 variants (**Table S1**). In addition, these findings correlate well with those of Frank et al. which demonstrated that none of the previously identified SARS-CoV-2 variants of concern containing point mutations in the BD Veritor System for Rapid Detection of SARS-CoV-2 antibody detecting antigen (Nucleocapsid protein) affected antibody recognition.^12^

Given the lack of SARS-CoV-2 specimens from asymptomatic individuals in our study, however, it would be beneficial to validate our findings using samples from that population. In addition, each WHO VOC/VOI classification contains several SARS-CoV-2 lineages and countless mutant strains and therefore, the ability to verify assay detection of every potential viral isolate was not feasible.

## CONCLUSION

The results of this study demonstrate the performance of the BD Veritor™ system for Rapid Detection of SARS-CoV-2 across circulating viral strains. However, since it is not possible to predict future mutations and genomic particularities of the SARS-CoV-2 virus, it is crucial that continued validation of the performance of SARS-CoV-2 assays be conducted, especially as new variants emerge, to support successful and optimal infection control strategies.

## Data Availability

Data produced in the present work are contained in the manuscript.

## ACKNOWLEDGMENTS

The authors thank Hélène Tanguay, Ph.D., MWC, of Becton, Dickinson and Company, BD Life Sciences – Integrated Diagnostic Solutions, for providing medical writing and editorial support. The authors are also grateful to the study participants who allowed this work to be performed.

## AUTHOR CONTRIBUTIONS

Karen Eckert: Supervision, Writing – Review and Editing; Sebastian Gutierrez: Formal analysis, Data curation, Writing – Review and Editing, Visualization, Project administration; Brittany Knight: Methodology, Formal analysis, Investigation, Writing – Review and Editing; Lauren Cooper: Data curation, Writing – Review and Editing, Supervision.

## SUPPLEMENTAL TABLE

**Table S1.** Variants detected using live, or heat inactivated isolates with the BD Veritor™ Plus System SARS-CoV-2 antigen test.

| WHO<br>Nomenclature | Variant | Reference Isolate Tested** |
| --- | --- | --- |
| **Alpha** | **B.1.1.7** | SARS-Related Coronavirus 2, Isolate USA/CA_CDC_5574/2020, BEI Resources Catalog No. NR-54011 |
| **Beta** | **B.1.351** | SARS-Related Coronavirus 2, Isolate hCoV-19/South Africa/KRISP-K005325/2020, BEI Resources Catalog No. NR-54009 |
| **Gamma** | **P.1** | SARS-Related Coronavirus 2, Isolate hCoV-19/Japan/TY7-503/2021, BEI Resources Catalog No. NR-54982 |
| **Kappa** | **B.1.617.1** | SARS-Related Coronavirus 2 Isolate hCoV-19/USA/CA-Stanford-15_S02/2021, BEI Resources Catalog No. NR-55486 |
| **Iota** | **B.1.526** | SARS-Related Coronavirus 2 Isolate hCoV-19/USA/NY-NP-DOH1/2021, BEI Resources Catalog No. NR-55359 |
| **Delta** | **B.1.617.2** | SARS-Related Coronavirus 2 Isolate hCoV-19/USA/PHC658/2021, BEI Resources Catalog No. NR-55611 |
| **Lambda** | **C.37** | U.S. NIH RADx Tech Team 6496 Screening Panel (Sept 23, 2021) |
| **Mu\*** | **B.1.621** | U.S. NIH RADx Tech Team 6496 Screening Panel (Sept 23, 2021) |
| **Omicron** | **B.1.1.529, BA.X**<br>(Not BA.2) | SARS-CoV-2, Isolate hCoV-19/USA/MD-HP20874/2021, BEI Resource Catalog No. NR-56461 |
| **Omicron** | **BA.2** | hCoV-19/USA/MD-HP24556-PIDGTQRIGP/2022 |
| **Omicron** | **BA.4** | hCoV-19/USA/MD-HP30386-PIDNBNVCCQ/2022 |
| **Omicron** | **BA.5.3.1** | hCoV-19/USA/MD-HP33394-PIDZONNRKD/2022 |

\* The Mu variant (B.1.621) contains the same characteristic T205I mutation as the Beta variant.
\*\* For each variant assessed, specific analytical reactivity data was generated using live or heat inactivated isolates on the BD Veritor™ SARS-CoV-2 Assay with BD Veritor™ Plus Analyzer. Commercially available nasal fluid from a single donor or multiple donors were pooled together in a 15ml conical tube to create the matrix used as a diluent. This matrix was screened in 3 replicates on the BD Veritor™ Plus Analyzer and confirmed to be negative which qualified its use in this study. Each dilution level was tested in at least three replicates to determine the tentative limit of detection or level of inclusivity. The tentative limit of detection was confirmed via testing in twenty replicates. Each replicate was analyzed with the BD Veritor Analyzer. Results from these studies have all confirmed detectability of variants by the BD Veritor™ Plus System SARS-CoV-2 antigen test.

